# Diagnostic clues and pitfalls in pontocerebellar hypoplasia type 2A

**DOI:** 10.1101/2025.06.10.25328274

**Authors:** Antonia Herrmann, Alice Kuhn, Maren Hackenberg, Julia Matilainen, Simone Mayer, Samuel Groeschel, Markus Uhl, Ingeborg Krägeloh-Mann, Wibke G. Janzarik

## Abstract

**Introduction:** Pontocerebellar hypoplasia type 2A (PCH2A) is a rare autosomal recessive neurodegenerative disease caused by a specific pathogenic variant in the TSEN54 gene (p.A307S). Affected children show early but initially unspecific symptoms, diagnosed primarily through postnatal MRI, with confirmation by genetic testing. This study examines the diagnostic process and key considerations for accurate diagnosis.

**Patients and Methods:** We retrospectively collected data from 65 children (33 female, 32 male) with genetically confirmed PCH2A as part of a Natural History Study. Data were gathered via parental questionnaires, interviews, and medical reports. The cohort was divided into two groups based on year of birth: children born before (n=30) and after (n=35) the identification of the pathogenic variant in 2008.

**Results:** Prenatally, in 4 of 21 cases with specialized ultrasound (gestational weeks 12-32), only unspecific cerebebellar abnormalities were reported. One fetal MRI (week 31) revealed clear cerebellar hypoplasia, in two others (week 21 and 31), slight cerebellar abnormalities were reported. Postnatal neurosonography often indicated disease features (26/54), later confirmed by MRI (62/63). Clinical symptoms appeared at a median age of 0 months (range 0-6 months), often initially suggesting acute rather than congenital issues. In the group born after 2008, median time from first symptoms to genetic confirmation was 5 months.

**Conclusion:** PCH2A presents early with nonspecific symptoms. Prenatal and postnatal ultrasound imaging can fail to detect the condition, with MRI being the gold standard for diagnosis. Over time, the diagnostic process, including genetic confirmation, has become faster.

**Highlights:**

- **Diagnostic Challenges**: Prenatal imaging fails to detect PCH2A
- **Postnatal Imaging**: MRI 92% diagnostic sensitive, ultrasound misinterpretations
- **Genetic testing impact**: Earlier PCH2A diagnosis in children born after 2008

## Introduction

Pontocerebellar hypoplasia (PCH) refers to a heterogeneous and expanding group of rare, predominantly neurodegenerative disorders inherited in an autosomal recessive manner. These conditions primarily affect the cerebellum and pons, though they may also involve supratentorial brain structures [1, 2]. The most common subtype, PCH2A, is a well-defined and homogeneous disorder caused by a homozygous pathogenic variant in the *TSEN54* gene (p.A307S), which probably alters tRNA processing and mRNA modification [3, 4, 5, 6].

Prenatal imaging in PCH2A if often reported as normal, although subtle abnormalities may occasionally be detected. Some reports describe a reduced transcerebellar diameter (TCD) on prenatal imaging, as well as mild clinical findings, such as head circumference at the lower limit of normal, polyhydramnios, or abnormal fetal movements [3, 7, 8, 9].

Postnatally, affected children typically present with symptoms within the first days of life [3, 7]. Newborns with PCH2A often show feeding difficulties, excessive crying, altered muscle tone, and episodes of drowsiness or restlessness [2, 3, 5]. A hallmark feature is progressive microcephaly, although head circumference at birth is often within the lower normal range [10, 11].

In infancy and early childhood, more specific symptoms emerge, including a dyskinetic movement disorder, global developmental delay, severe gastroesophageal reflux, episodic pain episodes with restlessness, and, later on, epileptic seizures [1, 3, 5, 12]. The overall life expectancy of children with PCH2A is significantly reduced [3, 11].

For families with a known history of PCH2A, prenatal genetic testing can provide a reliable diagnosis [13]. However, due to the autosomal recessive inheritance pattern, many parents are unaware of the genetic risk until a child is diagnosed with PCH2A. Postnatally, the diagnosis is typically suggested by MRI findings and confirmed through genetic testing [3, 7].

On postnatal MRI, the characteristic pontocerebellar hypoplasia often appears in a so-called *“dragonfly”* configuration, with relative sparing of the cerebellar vermis [14, 15]. Additional supratentorial abnormalities, such as cerebral atrophy, ventriculomegaly, and delayed myelination, have also been reported [7, 10, 14, 16, 17].

Differential diagnoses - clinically and radiologically - include other forms of PCH, Hoyeraal-Hreidarsson syndrome, congenital disorders of glycosylation, or infratentorial malformations such as Dandy-Walker syndrome, all of which can present with reduced cerebellar volume or be misinterpreted as such [1, 2, 14, 18]. Notably, brain lesions in extremely preterm infants can also mimic pontocerebellar hypoplasia [2, 19].

This retrospective study of 65 patients with genetically confirmed PCH2A aims to assess the diagnostic value and limitations of pre- and postnatal imaging and early clinical signs, with a focus on potential diagnostic pitfalls and misinterpretations on the way to the final diagnosis.

## Methods

Clinical data of PCH2A patients were retrospectively collected via digital questionnaires completed by parents. The study was approved by the Ethics Committee of the University of Freiburg (approval no. 20-1040), and the previous study was approved by the Ethics Committees of the University of Tübingen in 2012 (approval no. 105/2012BO2) and 2021 (approval no. 961/2020BO2). Written informed consent was obtained from all parents. This study specifically focused on pregnancy-related factors and diagnostic procedures. Not all parents answered every question, and some questions allowed for multiple responses; therefore, the reported percentages may not refer to the full cohort (N=65). In addition, medical reports and imaging data were reviewed to supplement the clinical information provided by the parents. Any discrepancies or missing data were addressed through follow-up telephone interviews with caregivers. For preterm infants, corrected age was used in the analysis of MRI data.

The study compared the time to suspected PCH and the time to definitive diagnosis between children born before and after 2008, the year the genetic basis of PCH2A was identified. After 2008, genetic testing became available to confirm the diagnosis of PCH2A [4].

Statistical analysis was conducted using SPSS and Excel, focusing on the calculation of medians, means, and the application of the Mann-Whitney U test to assess differences between the groups. Most of the results were analyzed descriptively and visualized using PowerPoint.

## Results

### Study Population

A total of 65 children with PCH2A were included in the study. Half of them (33 children) had previously participated in the initial Natural History Study (NHS) conducted 10 years earlier [3]. For 12 of these children, no follow-up data was available (one was still alive, four had passed away at the time of the first NHS, and seven were lost to follow-up). The participating families were from Germany, Switzerland, Austria, and Bulgaria. The gender distribution was even, with 33 girls (51%) and 32 boys (49%). The median age of the participants at the time of inclusion was 8.4 years, with a range of 7 months to 33 years. Of the 65 children, 42 (65%) were alive at the time of the survey, 16 (25%) had passed away, and survival status was unknown for 7 children (10%). Thirty children (46%) were born before 2008, while 35 children (54%) were born after. Nine sibling pairs were included in the study: seven pairs born before 2008, one pair born after 2008, and one pair with one child from each group.

### Cerebral Imaging

#### Prenatal Cerebral Imaging

Of the 65 participating children, 23 (35.4%) underwent specialized prenatal diagnostics using ultrasound (n=21) or MRI (n=3) (Figure 01). The median gestational age at the time of these examinations was 21 weeks (range: 12 to 32 weeks). Of the 23 cases with prenatal imaging, mild cerebellar abnormalities were noted in 7 cases (30.4%): in all 3 MRI cases and 4 of the 21 ultrasound examinations. These included findings such as “cerebellar hypoplasia,” “enlarged cisterna magna,” “suspected Dandy–Walker syndrome,” and “fluid around the cerebellum.” In two cases, fetal MRI was interpreted descriptively as showing nonspecific findings according to the available medical reports: an “enlarged posterior fossa without cerebellar hypoplasia” at 21 weeks of gestation, and “slender cerebellar hemispheres” at 31 weeks, respectively. In both cases, ultrasound examinations at 20–21 weeks showed normal transcerebellar diameters (TCDs). In the third case, a reduced TCD below the 5th percentile was reported in an ultrasound examination at 30 weeks, which was then confirmed by MRI at 31 and 34 weeks, with no increase in TCD between the two MRIs. Apart from imaging, additional specialized prenatal tests were performed in some cases, including amniotic fluid analysis (3/65), echocardiography, Doppler studies, and nuchal translucency measurements (each 2/65). In over half of the pregnancies (55.4%), no specialized investigations were performed.

**Figure 01.**
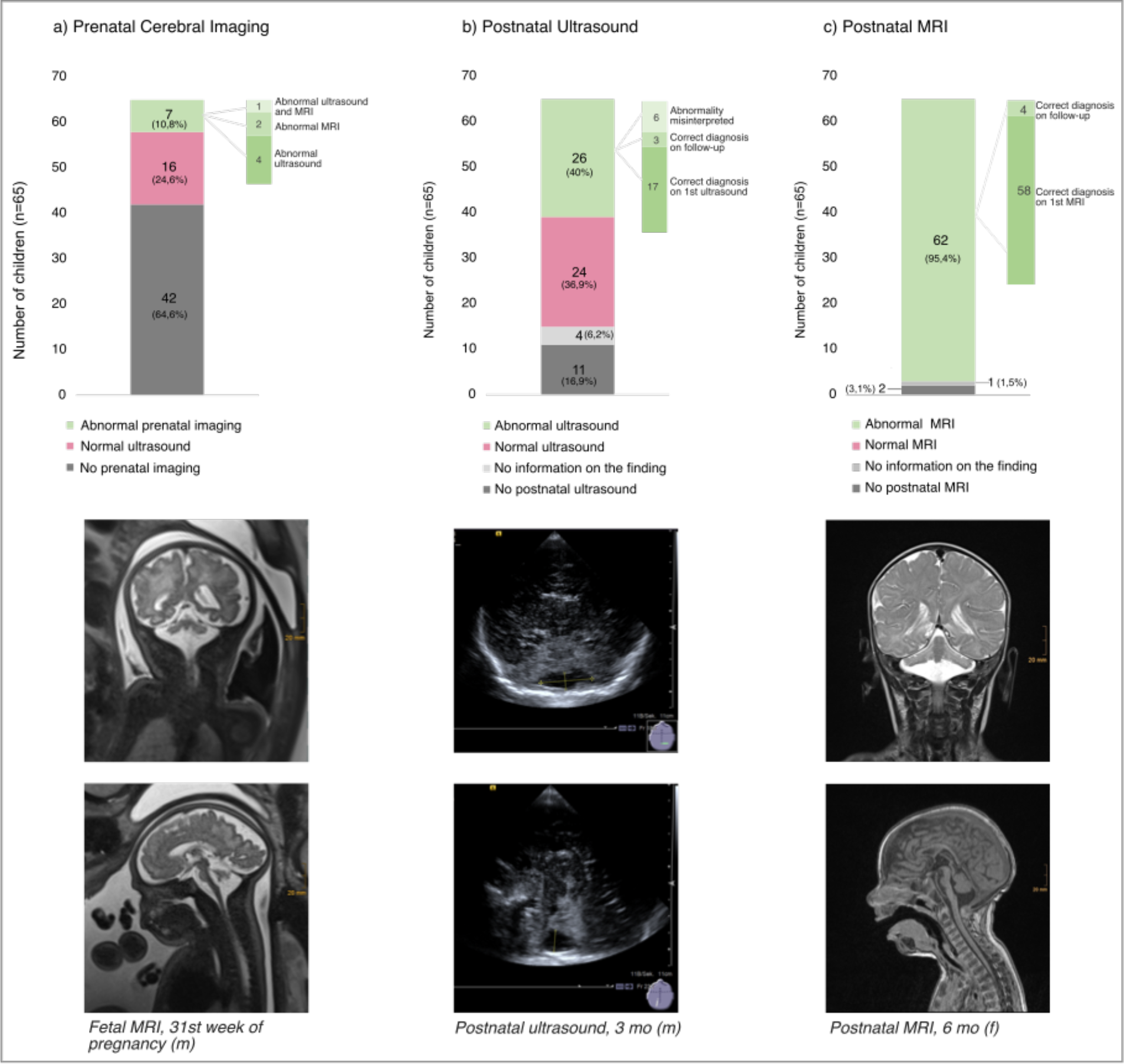
Cerebral Imaging Prenatal (a) and Postnatal (b+c). Top: Overview of all 65 participating children and their imaging. In grey: children who did not receive the corresponding imaging. Light grey: Children for whom no information on the findings was available. Red: Children for whom the respective imaging was classified as normal. Green: Children with a pathology in the imaging. These are further subdivided for a) Prenatal imaging: from dark to light: abnormal ultrasound, abnormal MRI, both entities abnormal. b) Postnatal ultrasound: from dark to light: correct diagnosis on first ultrasound, in follow-up, findings interpreted differently (megacisterna magna, cyst). c) Postnatal MRI: dark for correct diagnosis at first MRI and light for correct diagnosis during follow-up. Bottom: Example images of patients with PCH2A from our study.

#### Postnatal Ultrasound

Postnatal cranial ultrasound was performed in 54 out of 65 children (83.1%), with follow-up examinations in 20 children (37%) (Figure 01). The median age at the first ultrasound was 0 months, and the latest was at 12 months. Posterior fossa abnormalities were identified in nearly half of the children (26/54), with “cerebellar hypoplasia” or “enlarged cisterna magna” observed in 20 cases, primarily during the first examination. Six additional children had posterior cranial fossa abnormalities, initially misinterpreted as “cyst” (n=6) or “suspected Dandy-Walker syndrome” (n=3). Twelve children (22%) had normal ultrasound findings, and data was unavailable for four children. Other findings included difficult ultrasound conditions due to narrow or prematurely closed fontanelles (n=13), enlarged ventricles (n=11), and abnormal gyration patterns. Detailed findings are listed in Table 01.

**Table 01.** Findings in Postnatal Cerebral Ultrasound and MRI.

|  | Postnatal Ultrasound | Postnatal MRI |
| --- | --- | --- |
| Differential diagnosis/<br>misinterpretation | • Megacisterna Magna: 11/54 (20,4 %)<br>• Dandy-Walker: 3/54 (5,6 %)<br>• Cyst: 6/54 (11.1 %) | • Megacisterna Magna: 12/63 (19 %)<br>• Dandy-Walker: 5/63 (7.9 %)<br>• Blake-Pouch: 1/63 (1.6 %) |
| Additional findings | • Ventricular system: 11/54 (20.4 %)<br>• Narrow corpus callosum: 2/54 (3.7 %)<br>• Narrow fontanelle: 13/54 (24.1 %)<br>• Intraventricular hemorrhage: 3/54<br>• <u>Other</u>: Cyst in lateral ventricle/plexus (2/54), gyration anomaly (1/54) | • Ventricular system: 15/63 (23.8 %)<br>• Narrow corpus callosum: 10/63 (15.9 %)<br>• <u>Supratentorial</u>:<br>Volume reduction (7/63), delayed myelination (7/63), white matter anomalies (4/63), incomplete opercularization (3/63), gyration anomaly (2/63), SDH/SAB (1/63)<br>• <u>Other</u> (each 1/63):<br>Hemorrhage in posterior cranial fossa, edematous pons, arachnoid cyst, hemodynamic infarcts |

#### Postnatal MRI

All but two children underwent at least one MRI; 16 had follow-up MRIs, and seven underwent three or more scans (Figure 01). The median age at first MRI was 2.5 months. Pontocerebellar hypoplasia or abnormal cerebellar structure was identified in nearly all children (46/63), primarily on the initial scan (92.1%). In two cases, the first MRI was reported as normal, with the diagnosis established on follow-up imaging. Additionally, non-PCH-related abnormalities were frequently observed, including “ventricular enlargement” (15/63), “corpus callosum hypoplasia” (10/63), “delayed myelination” (7/63), and “thin white matter” (4/63). Dandy-Walker syndrome and Blake’s pouch cyst were noted in six children, representing important differential diagnoses. Table 01 provides detailed information on the MRI findings.

### Clinical Findings

#### Pregnancy

Approximately half of the pregnancies were reported as normal (30/65). The most common abnormality was unusual fetal movements, such as twitching or trembling (18/65), with either increased (8/65) or decreased (6/65) movement patterns. Polyhydramnios occurred in 12 pregnancies (18.5%), and oligohydramnios in one. Other findings included microcephaly (4 children) and intrauterine growth retardation (3 children). All pregnancy-related symptoms are visualized in Figure 02.

**Figure 02.**
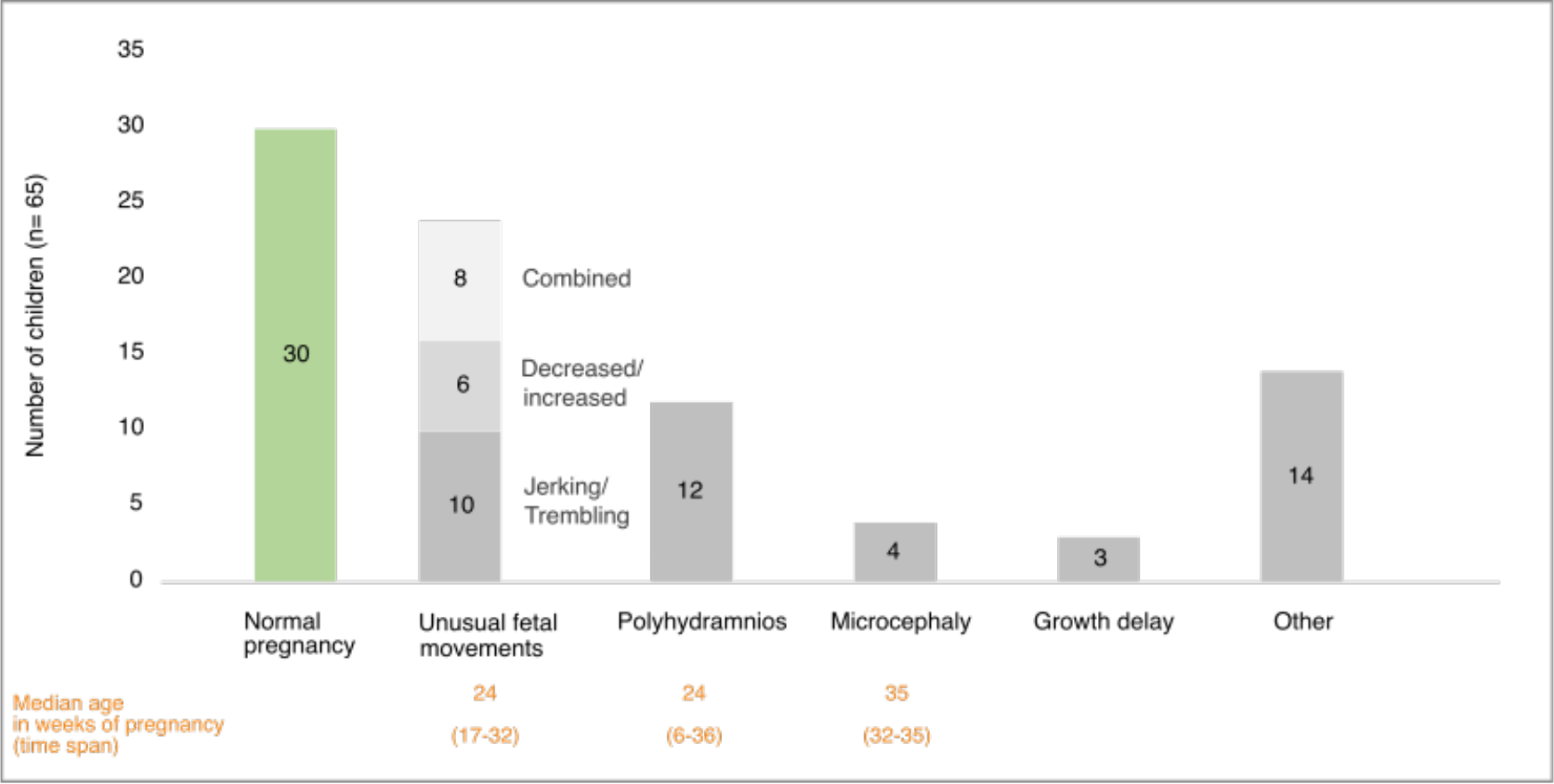
Abnormalities in Pregnancy and the Median Age of their Occurrence. Answers to the question of which symptoms the mothers observed during pregnancy. Multiple answers were possible. The left-hand bar (green) shows all children whose pregnancy was completely normal.

#### Birth

Most children were born full-term, with a median gestational age of 39 weeks and 4 days. Three children were born preterm, at 30+4, 36+0, and 36+4 weeks. APGAR scores at 5 and 10 minutes were mostly between 9 and 10. Umbilical artery pH values were normal in most children, though nine (17.3%) had acidotic values (≤7.2). Birth complications included shoulder dystocia (n=4), meconium-stained amniotic fluid (n=3), and breech presentation (n=2), as reported by parents. In four cases, parents stated that their children were born by caesarean section.

#### Neonatal Period

Hospital admission was required for 73.6% of children (39/53), typically within the first 72 hours of life. Interventions included nasogastric feeding tubes (42%), home monitoring (29%), and oxygen therapy (27%). Common neonatal issues included feeding difficulties (94%), restlessness (68%), and muscle hypertonia (66%). Further details on the neonatal period are shown in Figure 03.

**Figure 03.**
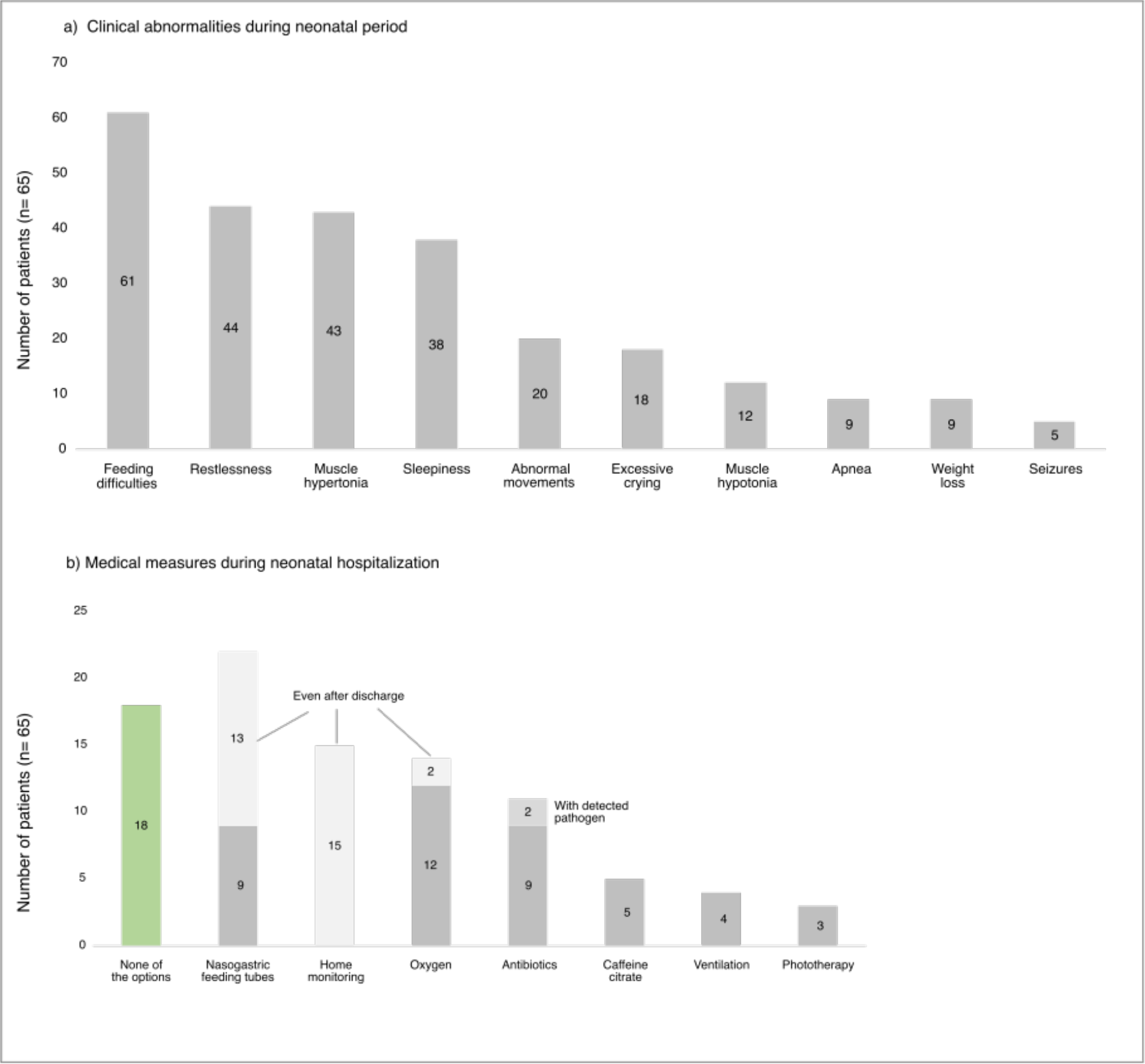
Symptoms and Therapeutic Measures during the Neonatal Period. a) Symptoms during the neonatal period, observed by parents and treating physicians, multiple answers possible. b) Therapeutic measures that were necessary for children who had to remain hospitalized after birth (n= 39), multiple answers possible.

#### First clinical Abnormalities observed by Parents and Pediatricians

Parents reported the earliest abnormalities within the first month of life (range 0–6 months). Feeding difficulties (n=52), vomiting (n=22), and developmental delays (n=28) were frequently noted as early signs. Other early observations included unusual movements, muscle hypertonia (n=35), hypotonia (n=9), myoclonies, hyperexcitability, and excessive crying. Additionally, 19 children were reported to have microcephaly, and seizures were among the first symptoms in 15 children. Pediatricians similarly noted feeding difficulties and abnormal cerebral ultrasound findings as the earliest signs, with a median age of 1 month (range 0–6 months). Further details are displayed in Figure 04.

**Figure 04.**
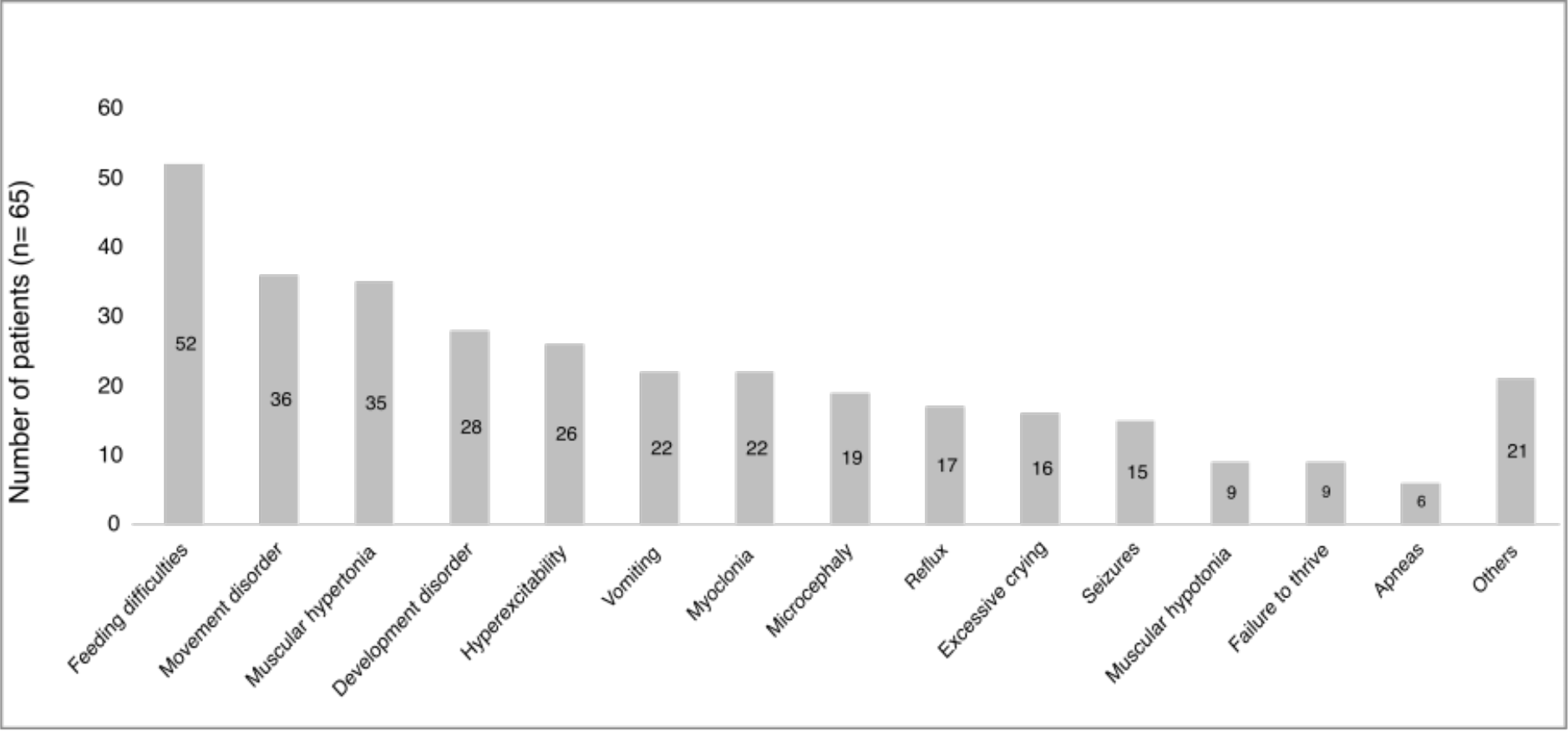
First Clinical Findings of PCH2A as observed by Parents. Multiple answers possible.

### Timeframe of Diagnostic Milestones

Children born before and after 2008 were compared regarding diagnostic milestones (see also Table 02). Parents were informed that PCH was suspected at a median age of 4 months. The difference between the two groups was not significant (p=0.13), with a median age of suspected diagnosis of 5 months (range: 0–364 months) for children born before 2008, and 4 months (range: 0–48 months) for those born after 2008. Genetic testing confirmed the diagnosis in 59 children (91%), with children born after 2008 being diagnosed at a median age of 8 months, compared to 4.75 years for those born before 2008. The time from first symptoms to genetic confirmation was significantly shorter for children born after 2008 (median 5 months) than for those born earlier (median 4.6 years). All median ages and quantiles are visualized in Figure 05. The results of the Mann-Whitney U test revealed a significant difference between the two groups only for genetic testing (U=70.5, Z=-5.399, p<0.001), with higher ranks observed in the pre-2008 group. This was also reflected in the time span from the first symptoms to the genetic test (U=61.5, Z=-5.538, p<0.001). The mean age at all other diagnostic steps did not significantly differ.

**Figure 05.**
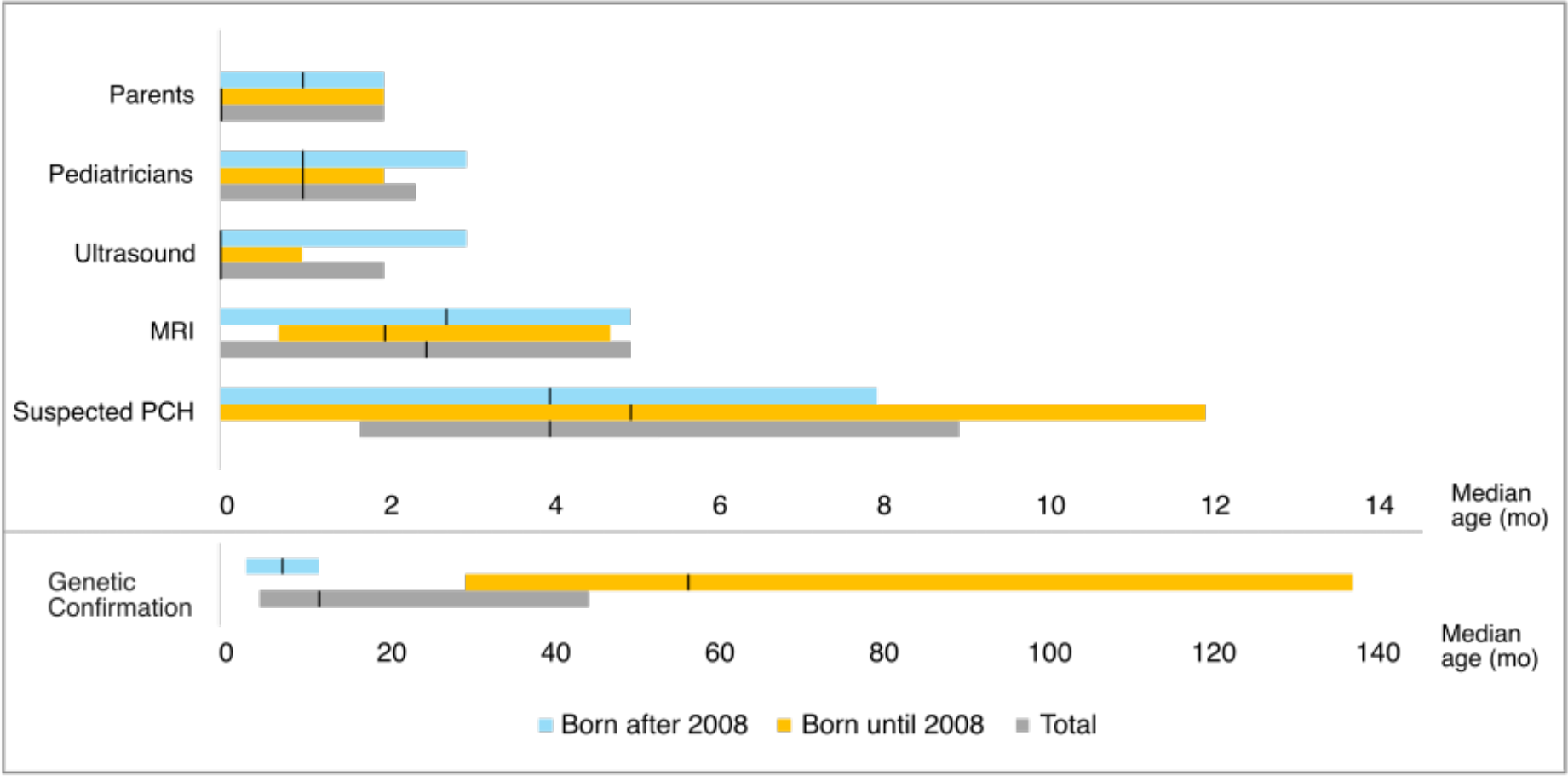
Timeline of Diagnostic Steps. Presentation of the various diagnostic steps from the first clinical abnormalities for the parents and pediatricians to the first ultrasound diagnosis, the first MRI and the expressed suspicion of PCH through to genetic confirmation. The median age of the children in months (black line) and the 25- and 75%-quantiles (bars) are shown, without including outliers in the illustration. A different scale was used for genetic confirmation (bottom). The color coding stands for the two groups separated by the year of birth up to and including 2008 (orange) and after 2008 (blue). In gray the total cohort.

**Table 02.** Median Age at Diagnostic Milestones.

|  | Median age in months (range) |  |  |
| --- | --- | --- | --- |
|  | Total (N=65) | Born until 2008 (n=30) | Born after 2008 (n=35) |
| Clinical abnormalities first observed by parents | 0 (0-6) | 0 (0-3,5) | 1 (0-6) |
| Clinical abnormalities first observed by pediatrician | 1 (0-6) | 1 (0-6) | 1 (0-6) |
| First ultrasound | 0 (0-12) | 0 (0-12) | 0 (0-5) |
| First MRI | 2,5 (0-146) | 2 (0-146) | 2,75 (0-24) |
| Suspected PCH | 4 (0-364) | 5 (0-364) | 4 (0-48) |
| Genetic confirmation | 12 (0-364) | 57 (4-364) | 8 (0-50) |
| Time interval between symptom onset and genetic diagnosis | 10 (0-364) | 55,5 (2-364) | 5 (0-148) |

## Discussion

### Prenatal Diagnosis of PCH2A

Prenatal diagnostic findings in PCH2A are rarely reported, and pregnancies are often described as uneventful [2, 7, 9, 13]. However, fetal MRI has been shown to be able to accurately diagnose PCH with some imaging features predicting classic PCH [8]. Although, as shown in our study, prenatal imaging findings of PCH2A can be detected prenatally, but often remain unrecognized. This is due not only to the limited spatial resolution of prenatal ultrasound in the posterior fossa and the general lack of awareness of this rare disease entity, but also to the physiological trajectory of cerebellar development, which accelerates in the third trimester - rendering deviations from the norm still subtle at the time of routine anomaly screening [20, 21].

The transcerebellar diameter (TCD) is a key marker in assessing cerebellar development and can be measured via both ultrasound and fetal MRI [8, 22–24]. TCD reduction has been described at birth in various PCH types, including PCH2A [8, 16, 25]. Jaillard et al. demonstrated in a heterogeneous cohort of fetuses with pontocerebellar hypoplasia, including three with PCH2A, that deviations from normal TCD values become more pronounced toward the end of gestation, coinciding with the accelerated cerebellar growth in the third trimester [8]. As the retrospective analysis identified 42 fetuses with decreased cerebellar biometry - only three of whom had PCH2A, the most common postnatal PCH subtype - the authors concluded that TSEN54-related PCH is rarely detected prenatally [8].

In our cohort, reported prenatal abnormalities of fetuses with PCH2A were generally vague and poorly quantified, particularly with regard to cerebellar hypoplasia. Retrospective analysis indicates that some of these unspecific findings may, in fact, reflect disease-related alterations. In at least one case, TCD measured in the third trimester was below the 5th percentile with no growth observed between 31 and 34 weeks of gestation, suggesting either impaired cerebellar development or early neurodegeneration.

Although fetal MRI offers improved visualization of posterior fossa structures compared to ultrasound, even MRI can fail to detect PCH2A in clinical routine due to the above mentioned reasons. Advanced MRI techniques such as fetal diffusion-weighted tractography, which could visualize the reduction of transverse pontocerebellar fibers - a pathognomonic finding in PCH2A - are not routinely available [13]. Furthermore, use of quantitative volumetric MRI analysis for prenatal MRI might also facilitate early diagnosis [10, 16].

Diagnosis is particularly challenging in the absence of family history. In our cohort, even in cases with known risk or polyhydramnios, prenatal MRI did not lead to an earlier (prenatal) diagnosis, and genetic testing was either declined or not initiated due to low clinical suspicion. Notably, none of the children born after 2008 - when genetic confirmation of PCH2A became available - underwent prenatal genetic testing, reflecting persistent under-recognition of this condition [4].

Due to the retrospective nature of our study, data on medically indicated terminations were not available. However, the high number of affected siblings born before 2008 suggests that increased access to and awareness of prenatal and preimplantation genetic testing could significantly improve early diagnosis and reproductive decision-making in at-risk families.

### Postnatal Imaging in PCH2A

Neurosonography is typically the first-line imaging modality in infants presenting with neurological concerns and was performed in the majority of our cases [26-28]. However, due to its limited ability to adequately visualize infratentorial structures, many findings remained vague or were misinterpreted, with terms such as “enlarged cisterna magna” frequently used. These limitations are consistent with previous reports [17].

Progressive microcephaly in patients with PCH2A typically emerges during infancy [10, 11]. In line with this, early closure of the anterior fontanelle was frequently observed in our cohort, likely reflecting impaired cerebral growth. This further restricted the acoustic window for ultrasound, complicating evaluation of posterior fossa structures [29, 30].

When cerebellar abnormalities are suspected, MRI is the modality of choice [26, 28]. In our study, nearly all children underwent brain MRI, with 92% demonstrating abnormal findings. However, interpretation was not always accurate. In two cases - including one preterm infant - initial MRI reports failed to detect cerebellar abnormalities, which only became apparent on follow-up scans. This suggests that in some cases, initial cerebellar changes are too subtle for early detection, reinforcing the value of serial imaging or quantification methods [2, 3, 10, 16].

Additional imaging findings included ventriculomegaly, callosal hypoplasia, delayed myelination, and reduced brain volume - none of which are specific for PCH2A [1, 7, 14]. The Evans Index was abnormal in all children older than 6.5 months, suggesting disproportionate ventricular enlargement [16]. Notably, Dandy-Walker malformation was suspected in five children, particularly in cases evaluated before 2008. This misinterpretation likely reflects limited awareness of PCH2A imaging features at the time and underscores the need for genetic confirmation when infratentorial abnormalities are identified.

### Clinical Indicators and Time to Diagnosis

Symptoms of PCH2A typically emerge in the neonatal period [2, 3, 7]. While some mothers retrospectively reported abnormalities already during pregnancy, these were often nonspecific. Most deliveries were uneventful; perinatal complications are more typical of more severe PCH types, such as PCH4 or PCH5 [2, 14]. Early progressive microcephaly is a key diagnostic clue of PCH2A [3, 11, 15]. Additional neurological abnormalities, such as altered tone with dyskinetic movements, feeding difficulties due to dysphagia, or persistent restlessness - should prompt further diagnostics in these infants.

In our cohort, the typical diagnostic sequence began with nonspecific symptoms in the first month, followed by neuroimaging at 2-3 months. A suspected diagnosis of PCH was usually reached within six months, with genetic confirmation around 12 months. For children born before 2008, diagnosis relied on clinical and radiological findings and was often delayed. Since then, time to diagnosis has decreased substantially. This underscores the value of early genetic testing in guiding care and family planning [31].

### Strengths and Limitations

This study benefits from a relatively large cohort of genetically confirmed PCH2A cases, which is considerable given the rarity of the disorder. The comprehensive collection of detailed clinical and imaging data allows for an in-depth phenotypic characterization of early clinical features and diagnostic steps. Additionally, the inclusion of a subgroup analysis comparing patients born before and after 2008 - the year when the genetic basis of PCH2A was identified - provides valuable insight into changes in diagnostic practices and highlights improvements in time to diagnosis.

However, several limitations must be acknowledged. The retrospective design inevitably leads to variability in data quality, including inconsistencies in medical documentation and questionnaire detail. Recall bias is particularly relevant, especially for older children, as parents may have difficulty accurately recalling early-life symptoms and events. Furthermore, the presence of previously diagnosed index cases in families with multiple affected children likely facilitated earlier recognition and diagnosis in younger siblings, introducing a potential ascertainment bias.

Other limitations include the predominantly European origin of our cohort, which may limit the generalizability of the findings to other populations. Prenatal findings may be underreported due to limited availability and use of advanced imaging and genetic testing in earlier years. Additionally, imaging protocols were not standardized across centers, possibly affecting the consistency of findings.

## Conclusion

Our findings highlight the diagnostic challenges of PCH2A, especially in the prenatal setting. Infratentorial abnormalities likely begin prenatally and may be visualized by MRI in late gestation; however, they are often subtle and can remain unrecognized when imaging is done early during gestation. In such cases, even mild cerebellar deviations should prompt consideration of a severe neurodevelopmental disorder such as PCH2A. Increased awareness of abnormal cerebellar trajectories in the third trimester, combined with careful imaging evaluation and genetic testing in families with a known risk, could substantially improve early detection and support informed reproductive planning.

Postnatally, affected children typically present with a distinct constellation of neurological symptoms. Brain MRI frequently reveals characteristic pontocerebellar abnormalities that can guide targeted genetic testing. Improved awareness of early clinical signs and imaging features can facilitate timely diagnosis, enable earlier genetic counseling, and support informed reproductive choices in at-risk families.

### Statement to the use of generative AI and AI-assisted technologies in scientific writing

During the preparation of this work, the authors used ChatGPT in order to improve the manuscript. After using this tool, the authors reviewed and edited the content as needed and take full responsibility for the content of the published article.

## Data Availability

All data produced in the present study are available upon reasonable request to the authors

## References

1. Barth PG. Pontocerebellar hypoplasies: an overview of a group of inherited neurodegenerative disorders with fetal onset. Brain Dev. 1993;15(6):411–22. doi:10.1016/0387-7604(93)90080-R.

2. Van Dijk T, Barth P, Baas F, Reneman L, Poll-The BT. Postnatal brain growth patterns in pontocerebellar hypoplasia. Neuropediatrics. 2021;52(3):163–9. doi:10.1055/s-0040-1716900.

3. Sánchez-Albisua I, Fröhlich S, Barth PG, Steinlin M, Krägeloh-Mann I. Natural course of pontocerebellar hypoplasia type 2A. Orphanet J Rare Dis. 2014;9:70. doi:10.1186/1750-1172-9-70.

4. Budde BS, Namavar Y, Barth PG, Poll-The BT, Nürnberg G, Becker C, et al. tRNA splicing endonuclease mutations cause pontocerebellar hypoplasia. Nat Genet. 2008;40(9):1113–8. doi:10.1038/ng.204.

5. Barth PG, Blennow G, Lenard HG, Begeer JH, Van Der Kley JM, Hanefeld F, et al. The syndrome of autosomal recessive pontocerebellar hypoplasia, microcephaly, and extrapyramidal dyskinesia (Pontocerebellar Hypoplasia Type 2): compiled data from 10 pedigrees. Neurology. 1995;45(2):311–7. doi:10.1212/WNL.45.2.311.

6. Hurtig JE, Steiger MA, Nagarajan VK, Li T, Chao TC, Tsai KL, et al. Comparative parallel analysis of RNA ends identifies mRNA substrates of a tRNA splicing endonuclease-initiated mRNA decay pathway. Proc Natl Acad Sci U S A. 2021;118(10):e2020429118. doi:10.1073/pnas.2020429118.

7. Steinlin M, Klein A, Haas-Lude K, Zafeiriou D, Strozzi S, Müller T, et al. Pontocerebellar hypoplasia type 2: variability in clinical and imaging findings. Eur J Paediatr Neurol. 2007;11(3):146–52.

8. Jaillard A, Valence S, Vande Perre S, Dhombres F, Héron D, Billette De Villemeur T, et al. Prenatal diagnosis of pontocerebellar hypoplasia with postnatal follow-up. Prenat Diagn. 2024;44(1):35–48. doi:10.1002/pd.6495.

9. Goasdoué P, Rodriguez D, Moutard ML, Robain O, Lalande G, Adamsbaum C. Pontoneocerebellar hypoplasia: de?ni7on of MR features. Pediatr Radiol. 2001;31(9):613–8. doi:10.1007/s002470100507.

10. Ekert K, Groeschel S, Sánchez-Albisua I, Fröhlich S, Dieckmann A, Engel C, et al. Brain morphometry in pontocerebellar hypoplasia type 2. Orphanet J Rare Dis. 2016;11(1):100. doi:10.1186/s13023-016-0481-4.

11. Kuhn A, Hackenberg M, Klauser AL, Herrmann A, Matilainen J, Mayer S, et al. Growth charts for pontocerebellar hypoplasia type 2A. Dev Med Child Neurol. 2025; (in press) doi:10.1101/2024.06.23.24307757

12. Janzarik WG, Krägeloh-Mann I, Langer T, van Buiren M, Schaefer HE, Gerner P. Spasmodic abdominal pain and other gastrointes7nal symptoms in Pontocerebellar Hypoplasia Type 2. Neuropediatrics. 2021;52(6):495–498. doi:10.1055/s-0041-1730445

13. Graham JM, Spencer AH, Grinberg I, Niesen CE, Plap LD, Maya M, et al. Molecular and neuroimaging ?ndings in pontocerebellar hypoplasia type 2 (PCH2): is prenatal diagnosis possible? Am J Med Genet A. 2010;152A(9):2268–76. doi:10.1002/ajmg.a.33579.

14. Uhl M, Pawlik H, Laubenberger J, Darge K, Baborie A, Korinthenberg R, et al. MR findings in pontocerebellar hypoplasia. Pediatr Radiol. 1998;28(7):547–51. doi:10.1007/s002470050410.

15. Namavar Y, Barth PG, Poll-The BT, Baas F. Classification, diagnosis and potential mechanisms in pontocerebellar hypoplasia. Orphanet J Rare Dis. 2011;6(1):50. doi:10.1186/1750-1172-6-50.

16. Pretzel P, Herrmann A, Kuhn A, Klauser AL, Matilainen J, Kellner E, et al. Brain morphometry and psychomotor development in children with PCH2A. Eur J Paediatr Neurol. 2025;45:1–9. doi:10.1016/j.ejpn.2025.04.004.

17. Pacheva IH, Todorov T, Ivanov I, Tartova D, Gaberova K, Todorova A, et al. TSEN54 gene-related pontocerebellar hypoplasia type 2 could mimic dyskinetic cerebral palsy with severe psychomotor retardation. Front Pediatr. 2018;6:1. doi:10.3389/fped.2018.00001.

18. Mahalingam H, Rangasami R, Seshadri S, Suresh I. Imaging spectrum of posterior fossa anomalies on foetal magnetic resonance imaging with an algorithmic approach to diagnosis. Pol J Radiol. 2021;86(1):183–94. doi:10.5114/pjr.2021.105014.

19. Messerschmidt A, Brugger PC, Boltshauser E, Zoder G, Sterniste W, Birnbacher R, et al. Disruption of cerebellar development: potential complication of extreme prematurity. 2005.

20. Volpe JJ. Cerebellum of the premature infant: rapidly developing, vulnerable, clinically important. J Child Neurol. 2009;24(9):1085–104. doi:10.1177/0883073809338067.

21. Koning IV, Dudink J, Groenenberg IAL, Willemsen SP, Reiss IKM, Steegers-Theunissen RPM. Prenatal cerebellar growth trajectories and the impact of periconcep7onal maternal and fetal factors. Hum Reprod. 2017;32(6):1230–1237. doi:10.1093/humrep/dex079

22. Tich SN, Anderson PJ, Shimony JS, Hunt RW, Doyle LW, Inder TE. A novel quantitative simple brain metric using MR imaging for preterm infants. AJNR Am J Neuroradiol. 2009;30(1):125–31. doi:10.3174/ajnr.A1309.

23. Torres HR, Morais P, Oliveira B, Birdir C, Rüdiger M, Fonseca JC, et al. A review of image processing methods for fetal head and brain analysis in ultrasound images. Comput Methods Programs Biomed. 2022;215:106629. doi:10.1016/j.cmpb.2022.106629.

24. Pogledic I, Mankad K, Severino M, Lerman-Sagie T, Jakab A, Hadi E, et al. Prenatal assessment of brain malformations on neuroimaging: an expert panel review. Brain. 2024;147(12):3982–4002. doi:10.1093/brain/awae253.

25. Patel MS, Becker LE, Toi A, Armstrong DL, Chitayat D. Severe, fetal-onset form of olivopontocerebellar hypoplasia in three sibs: PCH type 5? Am J Med Genet A. 2006;140A(6):594–603. doi:10.1002/ajmg.a.31095.

26. Van Wezel-Meijler G, Steggerda SJ, Leijser LM. Cranial ultrasonography in neonates: role and limitations. Semin Perinatol. 2010;34(1):28–38. doi:10.1053/j.semperi.2009.10.002.

27. Weise J, Heckmann M, Bahlmann H, Ittermann T, Allenberg H, Domanski G, et al. Analyses of pathological cranial ultrasound findings in neonates that fall outside recent indication guidelines: results of a population-based birth cohort: survey of neonates in Pommerania (SNiP-study). BMC Pediatr. 2019;19(1):476. doi:10.1186/s12887-019-1843-6.

28. AIUM Practice Parameter for the Performance of Neurosonography in Neonates and Infants. J Ultrasound Med. 2020;39(5). doi:10.1002/jum.15264.

29. Kiesler J, Ricer R. The abnormal fontanel. Am Fam Physician. 2003;67(12):2547–52.

30. Oumer M, Tazebew A, Alemayehu M. Anterior fontanel size among term newborns: a systematic review and meta-analysis. Public Health Rev. 2021;42:1604044. doi:10.3389/phrs.2021.1604044.

31. Ammann-Schnell L, Groeschel S, Kehrer C, Fröhlich S, Krägeloh-Mann I. The impact of severe rare chronic neurological disease in childhood on the quality of life of families - a study on MLD and PCH2. Orphanet J Rare Dis. 2021;16:211. doi:10.1186/s13023-021-01828-y.

